# Atypical Presentations of COVID-19 in Care Home Residents presenting to Secondary Care: A UK Single Centre Study

**DOI:** 10.1101/2020.07.07.20148148

**Authors:** Mark James Rawle, Deborah Lee Bertfield, Simon Edward Brill

**Author notes:** Corresponding author: Dr Mark James Rawle, Department of Geriatric Medicine, Barnet Hospital, Wellhouse Lane, E5N 3DJ.

## Abstract

**Purpose:** The United Kingdom (UK) care home population has experienced high mortality during the COVID-19 pandemic. Atypical presentations of COVID-19 are being reported in older adults and may pose difficulties for early isolation and treatment, particularly in institutional care settings. We aimed to characterise the presenting symptoms and associated mortality of COVID-19 in older adults, with a focus on care home residents and older adults living in the community.

**Methods:** This was a retrospective cohort study of consecutive inpatients over 80 years old hospitalised with PCR confirmed COVID-19 between 10^th^ March 2020 and 8^th^ April 2020. Symptoms at presentation, including those associated with frailty, were analysed. Differences between community dwelling and care home residents, and associations with mortality, were assessed using between-group comparisons and logistic regression.

**Results:** Care home residents were less likely to experience cough (46.9% vs 72.9%, p=0.002) but more likely to present with delirium (51.6% vs 31.4%, p=0.018), particularly hypoactive delirium (40.6% vs 24.3%, p=0.043). Mortality was more likely in the very frail (OR 1.25, 95% CI 1.00, 1.58, p=0.049) and those presenting with anorexia (OR 3.20, 95% CI 1.21, 10.09, p=0.028). There were no differences in either mortality or length of stay between those admitted from care homes and community dwelling older adults.

**Conclusion:** COVID-19 in those over 80 does not always present with typical symptoms, particularly in those admitted from institutional care. These individuals have a reduced incidence of cough and increased hypoactive delirium. Individuals presenting atypically, especially with anorexia, have higher mortality.

## 1. Introduction

In January 2020, the first laboratory-confirmed case of Coronavirus disease of 2019 (COVID-19), a febrile respiratory illness caused by a novel pathogenic strain of coronavirus, SARS-coronavirus-2 (SARS-CoV-2) [1] was reported in the United Kingdom (UK). As of 2^nd^ June 2020, there have been 276,332 confirmed cases and 39,045 deaths nationally [2], with potentially as many of a third of these deaths occurring in care home residents [3].

The typical symptoms of COVID-19 are predominantly respiratory (cough, sputum, sore throat, runny nose, ear pain, wheeze, and chest pain), systemic (myalgia, joint pain and fatigue) and enteric (abdominal pain, vomiting and diarrhoea) [4]. Due to age related physiological and immunological changes, older people commonly present without classic symptoms [5]. Data are emerging that presentation with COVID-19 in this cohort may also be atypical, potentially with geriatric syndromes including falls, delirium and anorexia [6-11].

Atypical presentation is of particular concern for residential and nursing home residents. Evidence exists that transmission within nursing homes is rapid due to difficulties in identifying new COVID-19 infections [12]. Isolation of suspected cases is critical to protect staff and other residents [13], but requires understanding of the characteristics of COVID-19 in this population.

Barnet is the most populous of London’s boroughs with an estimated population of 402,700 residents and approximately 16,800 people over the age of 80. There are more than 100 care homes within the borough [14]. Barnet Hospital is a general suburban hospital with 440 beds, and was affected by a high volume of COVID-19 cases early in the course of the UK pandemic [15]. Using admission data from this period we set out to a) characterise the presenting symptoms in older adults admitted with COVID-19, b) determine whether care home residents exhibited different presentations to the general older population, and c) examine the mortality associations with typical and atypical presentations. We hypothesised that atypical presentation would be more common in care home residents and associated with increased mortality.

## 2. Methods

### 2.1 Study design, setting and population

Patients aged 80 or over admitted to Barnet Hospital with COVID-19 confirmed on consecutive polymerase chain reaction (PCR) testing for SARS-CoV2 were included for analysis. Individuals who had continuously been an inpatient for 14 days beforehand were excluded on the basis of having acquired COVID-19 in hospital and therefore unrelated symptoms on admission. Patients with a clinical diagnosis of COVID-19 without PCR confirmation were not included. Subgroups were defined within this population based on their care needs prior to admission, including carers at home and institutional care. Data were collected retrospectively from the electronic patient record. Further information is available here [15].

### 2.2 Ascertainment of key variables

Standardised data were collected on demographic features, ethnicity, length of stay and the presence of co-morbidities (prior diagnosis of cardiac disease, hypertension, diabetes, respiratory disease, immunosuppression and dementia). Frailty was determined by the Clinical Frailty Scale (CFS) [16], assessed contemporaneously on admission and retrospectively validated independently by two consultant geriatricians (MJR & DLB).

Symptoms were assessed using electronic health records, and were taken from the day of admission only. Fever was defined as a temperature > 37.8 Celsius. Medical records were reviewed for the specific ‘geriatric syndromes’ of falls, delirium and anorexia by two consultant geriatricians (MJR & DLB). Delirium was categorised as either hypoactive or hyperactive based on presenting symptoms, and 4 ‘A’s Test [17] was used where available from initial clerking. Individuals were defined as having a fully atypical presentation if cough, fever and dyspnoea were absent at presentation.

The primary outcome assessed was death versus discharge from hospital at the end of the hospital episode. All included patients had reached this endpoint by the time of analysis.

### 2.3 Statistical analyses

Demographic and clinical characteristics of the cohort were described using measures of central tendency and variability to explore differences between residential status (community dwelling versus care home residents). Pearson’s chi-square and Fisher’s exact tests were used to test for relationships between categorical outcome variables and residential category. For relationships between continuous variables and residential category, the Wilcoxon signed-rank test was used.

We used univariable logistic regression to test the association between mortality and presenting symptoms. A series of logistic regressions were run to assess independent associations between the covariates and mortality. Any that were identified as statistically or clinically significant confounders (p<0.1), through regression analyses, were sequentially adjusted for in the final multivariable regression model for this association, with backwards elimination to remove variables one at a time until only statistically significant (p<0.05) variables remained in the model. All analyses were performed using R on a complete-case basis, including only participants with full data on delirium status and medication. All analysis was performed using R Statistics version 3.6.3.

### 2.4 Ethical approval

The data presented here were collected during routine clinical practice and formal Research Ethics Committee review was not required. Support for the study was given by the chair of the Trust Clinical Ethics Committee.

## 3. Results

### 3.1 Participant eligibility

150 individuals over the age of 80 admitted to Barnet Hospital with SARS-CoV2 confirmed on PCR between 10^th^ March 2020 and 8^th^ April 2020 were included. Of these individuals, 16 were excluded on the basis of having hospital acquired COVID-19. The remaining 134 individuals had complete data for all analysed variables. [Figure 1]

**Figure 1.**
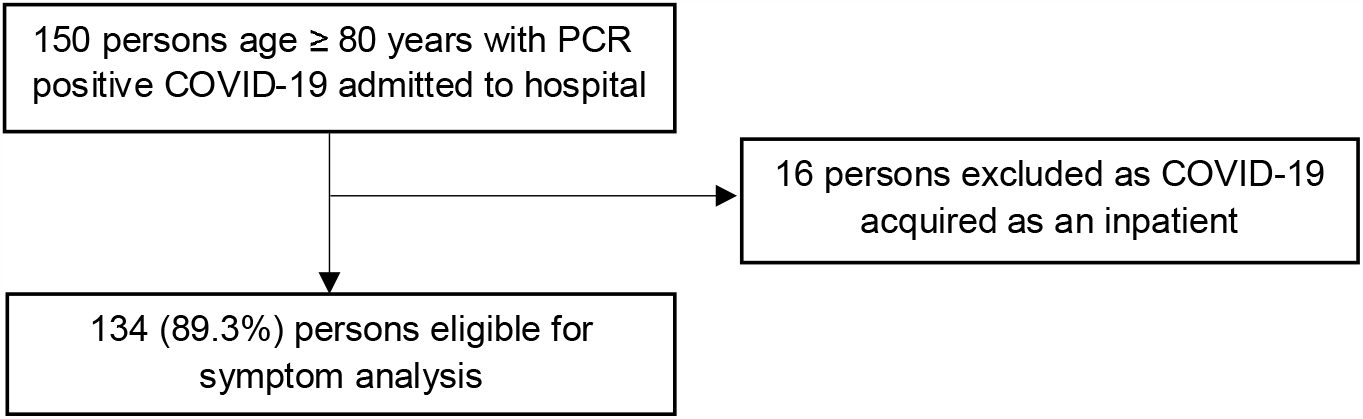
Study flowchart showing participant eligibility and exclusion process

**Figure 2.**
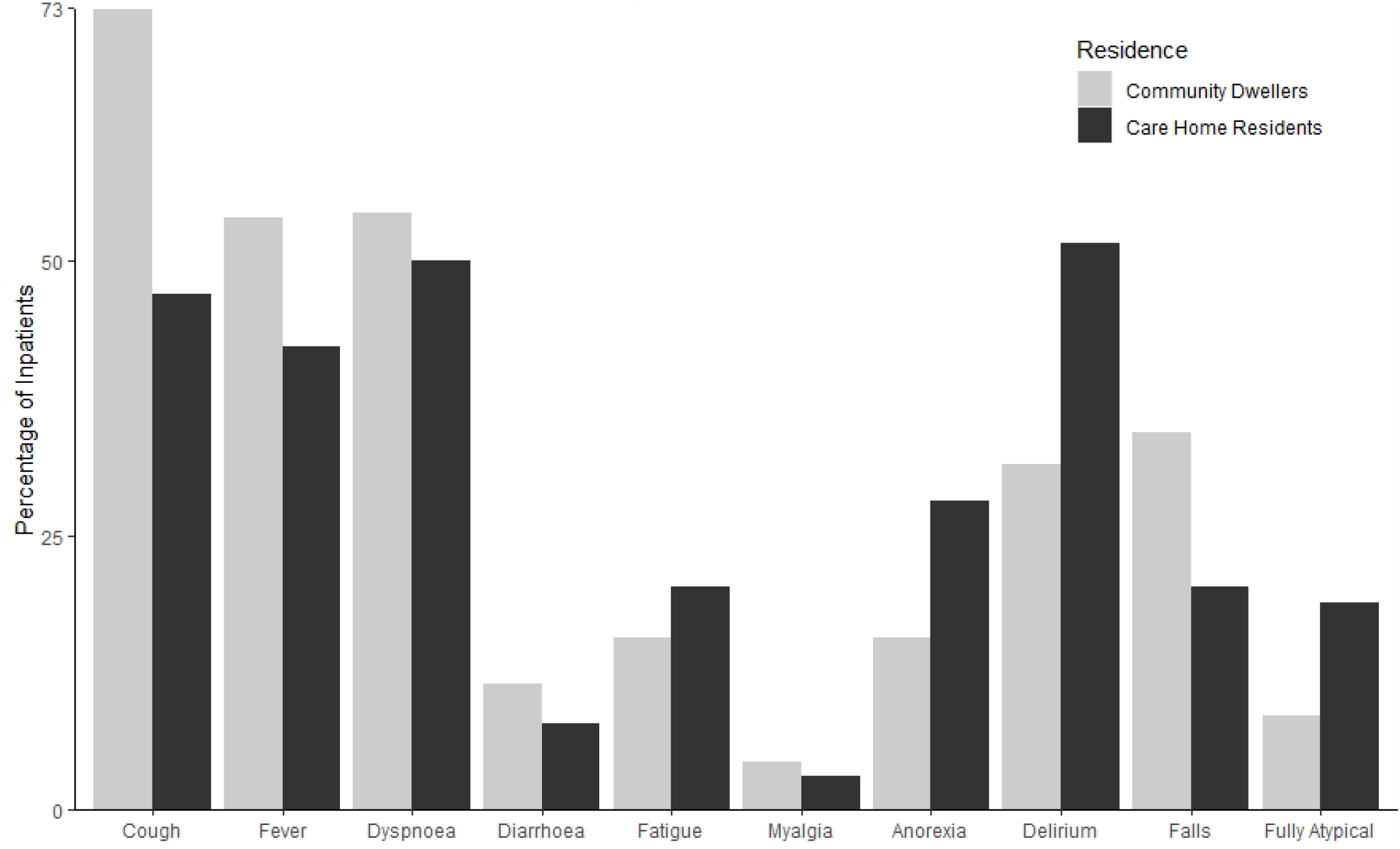
Bar plot of COVID-19 presenting symptoms by residential status

### 3.2 Cohort and clinical characteristics

The median age was 86 years, and most participants were white (76.1%, n=102). There was a slight male predominance (54.5%, n=73) and a nearly equal split between residential status (52.2% community dwelling participants, n=70). All CFS categories were represented with mild and moderate frailty (CFS 5 & 6) most prevalent. Most participants had one or more comorbidities, with hypertension (55.2%, n=74), cardiac disease (56.7%, n=76) and diabetes mellitus (29.1%, n=39) most common.

For those who reported symptoms, cough remained the most prevalent symptom (60.4%, n=81) with dyspnoea (52.2%, n=70) and fever (47.8%, n=64) also prevalent in those admitted. Atypical presenting symptoms were pronounced in older patients, with a high prevalence of delirium (41%, n=55) particularly hypoactive (32.1%, n=43), falls (27.6%, n=37), anorexia (21.2%, n=29) and fatigue (17.9%, n=24) reported. Just over a tenth of all patients presented fully atypically without fever, cough or dyspnoea (13.4%, n=18). The overall mortality was high (64.9%, n=87) with a median length of stay of 11 days for those that survived.

When compared to community dwelling older adults, care home residents admitted with COVID-19 tended to be older (median age 88.5 vs 85, p<0.001) and have higher frailty as indicated by the CFS (p<0.001). Both groups had a similar gender and ethnicity distribution, and a similar prevalence of comorbidities, although dementia was more prevalent in care home residents (37.5% vs 15.7%, p=0.004).Care home residents were less likely to present with cough (46.9% vs 72.9%, p=0.002) and more likely to present with delirium (51.6% vs 31.4%, p=0.018) particularly hypoactive delirium (40.6% vs 24.3%, p=0.043). There was a suggestion that fully atypical presentation was more common in care home residents, though total numbers were small, limiting significance (18.8% vs 8.6%, p=0.084). There was no notable difference in either mortality or length of stay between these two groups.

Full sample characteristics are provided in Table 1.

**Table 1.** Sample Characteristics by Residential Status. Count and percentages, accompanied by Pearson chi-square tests (or Fisher’s exact) to assess associations between variables by residential status, are presented for categorical variables. Median and interquartile range, with Kruskal-Wallis tests to assess differences in group medians, are presented for all continuous variables due to skewed distribution. Clinical frailty scale treated as continuous. *n* = number of participants, IQR = Inter Quartile Range, * = significant at p < 0.05.

| Variables | Total n (%) | Residential Status |  | p-value |
| --- | --- | --- | --- | --- |
|  |  | Community | Care Home |  |
| n (%) | 134 (100%) | 70 (52.2%) | 64 (47.8%) |  |
| Median age (IQR) | 86 (7.6) | 85 (6) | 88.5 (7) | **<0.001\*** |
| Male gender | 73 (54.5%) | 39 (55.7%) | 34 (53.1%) | 0.764 |
| Ethnicity |  |  |  |  |
| *White* | 102 (76.1%) | 49 (70.0%) | 53 (82.8%) |  |
| *Black* | 4 (3.0%) | 4 (5.7%) | - |  |
| *Asian* | 11 (8.2%) | 5 (7.1%) | 6 (9.4%) |  |
| *Other* | 17 (12.7%) | 12 (17.1%) | 5 (7.8%) | 0.071 |
| Clinical Frailty Scale |  |  |  |  |
| 1 | 3 (2.2%) | 3 (4.3%) | - |  |
| 2 | 6 (4.5%) | 6 (8.6%) | - |  |
| 3 | 13 (9.7%) | 13 (18.6%) | - |  |
| 4 | 14 (10.4%) | 14 (20.0%) | - |  |
| 5 | 35 (26.1%) | 20 (28.6%) | 15 (23.4%) |  |
| 6 | 36 (26.9%) | 9 (12.9%) | 27 (42.4%) |  |
| 7 | 20 (14.9%) | 3 (4.3%) | 17 (26.6%) |  |
| 8 | 6 (4.5%) | 1 (1.4%) | 5 (7.8%) |  |
| 9 | 1 (0.7%) | 1 (1.4%) | - | **<0.001\*** |
| Comorbidities |  |  |  |  |
| *Hypertension* | 74 (55.2%) | 35 (50.0%) | 39 (60.9%) | 0.203 |
| *Diabetes Mellitus* | 39 (29.1%) | 20 (28.6%) | 19 (29.7%) | 0.576 |
| *Cardiac Disease* | 76 (56.7%) | 38 (54.3%) | 38 (59.4%) | 0.553 |
| *Respiratory Disease* | 23 (17.2%) | 15 (21.4%) | 8 (12.5%) | 0.171 |
| *Immunosuppression* | 7 (5.2%) | 2 (2.9%) | 5 (7.8%) | 0.198 |
| *Dementia* | 35 (26.1%) | 11 (15.7%) | 24 (37.5%) | **0.004\*** |
| Presenting Symptoms |  |  |  |  |
| *Cough* | 81 (60.4%) | 51 (72.9%) | 30 (46.9%) | **0.002\*** |
| *Fever* | 64 (47.8%) | 37 (53.9%) | 27 (42.2%) | 0.217 |
| *Dyspnoea* | 70 (52.2%) | 38 (54.3%) | 32 (50.0%) | 0.620 |
| *Diarrhoea* | 13 (9.7%) | 8 (11.4%) | 5 (7.8%) | 0.488 |
| *Fatigue* | 24 (17.9%) | 11 (15.7%) | 13 (20.3%) | 0.488 |
| *Myalgia* | 5 (3.7%) | 3 (4.3%) | 2 (3.1%) | 0.723 |
| *Anorexia* | 29 (21.2%) | 11 (15.7%) | 18 (28.1%) | 0.081 |
| *Delirium* | 55 (41.0%) | 22 (31.4%) | 33 (51.6%) | **0.018\*** |
| *(hypoactive subtype)* | 43 (32.1%) | 17 (24.3%) | 26 (40.6%) | **0.043\*** |
| *(hyperactive subtype)* | 13 (9.7%) | 6 (8.6%) | 7 (10.9%) | 0.644 |
| *Falls* | 37 (27.6%) | 24 (34.3%) | 13 (20.3%) | 0.071 |
| Atypical Presentation | 18 (13.4%) | 6 (8.6%) | 12 (18.8%) | 0.084 |
| Mortality | 87 (64.9%) | 45 (64.3%) | 42 (65.6%) | 0.871 |
| Median Survivor Length of Stay (IQR) | 11 (7) | 8 (8) | 7.5 (8.3) | 0.165 |

### 3.3 Associations with mortality

Individuals were noted to be more likely to die when presenting with a higher CFS score (Odds Ratio (OR) 1.25, 95% CI 1.00, 1.58, p=0.049) or when presenting with anorexia (OR 3.20, 95% CI 1.21, 10.09, p=0.028). Those presenting with falls were less likely to die (OR 0.45, 95% CI 0.21, 0.98, p=0.044). No other assessed demographic factors (age, gender, ethnicity, comorbidities) were associated with an altered mortality rate, nor were any other presenting symptoms.

These trends were enhanced for individuals residing in care homes, with higher CFS being more strongly associated with increased mortality (OR 2.94, 95% CI 1.47, 6.66, p=0.005), along with anorexia (OR 6.15, 95% CI 1.51, 41.83, p=0.024) and the presence of pre-existing dementia (OR 4.09, 95% CI 1.27, 16.03, p=0.026). Individuals presenting from a care home with cough had a lower association with mortality (OR 0.26, 95% CI 0.08, 0.75, p=0.016). For care home residents, falls did not have any association with mortality. Individuals presenting atypically from care homes showed the suggestion of a greater association with mortality (OR 7.45, 95% CI 1.3, 141.36, p=0.063). Full univariable associations with mortality are presented in Table 2.

**Table 2.** Univariable Associations with Mortality by Residential Status. Statistics obtained through a series of logistic or linear regressions where appropriate. p values indicate significance of association between outcome and (level of) covariate. Odds s rounded to 2 decimal places. OR = odds ratio, 95% CI = 95% confidence interval, n = number of participants, ref = reference group, * = significant at p < 0.05.

| Mortality | Total Sample (n=134)<br>OR (95% CI) p-value | Residential Status |  |
| --- | --- | --- | --- |
|  |  | Community (n=70)<br>OR (95% CI) p-value | Care Home (n=64)<br>OR (95% CI) p-value |
| Residential Status (admitted from home) | 0.94 (0.46, 1.92) p=0.871 | - | - |
| Age (per additional year) | 1.03 (0.96, 1.10) p=0.466 | 1.02 (0.91, 1.14) p=0.752 | 1.04 (0.94, 1.15) p=0.498 |
| Male gender | 1.61 (0.79, 3.31) p=0.191 | 1.63 (0.61, 4.41) p=0.334 | 1.60 (0.57, 4.60) p=0.375 |
| Ethnicity |  |  |  |
| White | ref | ref | ref |
| Black | 0.57 (0.07, 4.90) p=0.581 | 0.63 (0.07, 5.64) p=0.661 | - |
| Asian | 1.00 (0.28, 4.01) p=0.995 | 2.53 (0.34, 51.57) p=0.421 | 0.51 (0.09, 3.02) p=0.443 |
| Other | 1.85 (0.60, 6.94) p=0.311 | 1.90 (0.49, 9.38) p=0.881 | 2.06 (0.28, 41.84) p=0.532 |
| Clinical Frailty Scale (per additional point) | 1.25 (1.00, 1.58) **p=0.049\*** | 1.18 (0.87, 1.64) p=0.291 | 2.94 (1.47, 6.66) **p=0.005\*** |
| Comorbidities |  |  |  |
| Hypertension | 1.14 (0.56, 2.32) p=0.728 | 1.46 (0.55, 3.95) p=0.455 | 0.84 (0.28, 2.41) p=0.749 |
| Diabetes Mellitus | 1.12 (0.51, 2.50) p=0.787 | 1.43 (0.48, 4.62) p=0.529 | 0.86 (0.28, 2.72) p=0.787 |
| Cardiac Disease | 1.42 (0.70, 2.92) p=0.333 | 1.15 (0.43, 3.09) p=0.775 | 1.8 (0.631, 5.20) p=0.271 |
| Respiratory Disease | 1.29 (0.50, 3.59) p=0.609 | 1.14 (0.35, 4.10) p=0.828 | 1.67 (0.35, 12.11) p=0.554 |
| Immunosuppression | 0.70 (0.15, 3.72) p=0.659 | 0.55 (0.02, 14.21) p=0.673 | 0.77 (0.12, 6.20) p=0.783 |
| Dementia | 2.19 (0.94, 5.62) p=0.082 | 0.97 (0.26, 4.05) p=0.961 | 4.09 (1.27, 16.03) **p=0.026\*** |
| Presenting Symptoms |  |  |  |
| Cough | 0.60 (0.28, 1.26) p=0.186 | 1.46 (0.48, 4.29) p=0.497 | 0.26 (0.08, 0.75) **p=0.016\*** |
| Fever | 0.93 (0.46, 1.90) p=0.841 | 0.82 (0.30, 2.19) p=0.695 | 1.08 (0.38, 3.15) p=0.881 |
| Dyspnoea | 1.40 (0.69, 2.87) p=0.356 | 1.91 (0.71, 5.23) p=0.200 | 1.00 (0.35, 2.83) p=1.000 |
| Diarrhoea | 3.26 (0.83, 21.66) p=0.136 | 4.42 (0.72, 85.35) p=0.177 | 2.21 (0.30, 44.75) p=0.491 |
| Fatigue | 0.71 (0.29, 1.79) p=0.456 | 0.97 (0.26, 4.05) p=0.961 | 0.53 (0.15, 1.90) p=0.321 |
| Myalgia | 0.35 (0.04, 2.16) p=0.253 | 1.12 (0.10, 24.76) p=0.930 | 0.00 (0.00, 0.00) p=0.992 |
| Anorexia | 3.20 (1.21, 10.09) **p=0.028\*** | 1.59 (0.41, 7.81) p=0.527 | 6.15 (1.51, 41.83) **p=0.024\*** |
| Delirium | 1.82 (0.87, 3.89) p=0.116 | 1.29 (0.45, 3.92) p=0.646 | 2.57 (0.90, 7.74) p=0.082 |
| (hypoactive subtype) | 1.90 (0.87, 4.39) p=0.116 | 1.46 (0.46, 5.13) p=0.534 | 2.42 (0.82, 7.88) p=0.120 |
| (hyperactive subtype) | 1.24 (0.38, 4.79) p=0.732 | 1.12 (0.20, 8.54) p=0.899 | 1.35 (0.26, 10.03) p=0.733 |
| Falls | 0.45 (0.21, 0.98) **p=0.044\*** | 0.52 (0.184, 1.44) p=0.205 | 0.36 (0.10, 1.24) p=0.105 |
| Atypical Presentation | 3.06 (0.94, 13.73) p=0.091 | 1.12 (0.20, 8.54) p=0.899 | 7.45 (1.3, 141.36) p=0.063 |

For care home residents these associations persisted when combined in a multivariable model. Our multivariable model for care home residents is presented in Table 3.

**Table 3.**
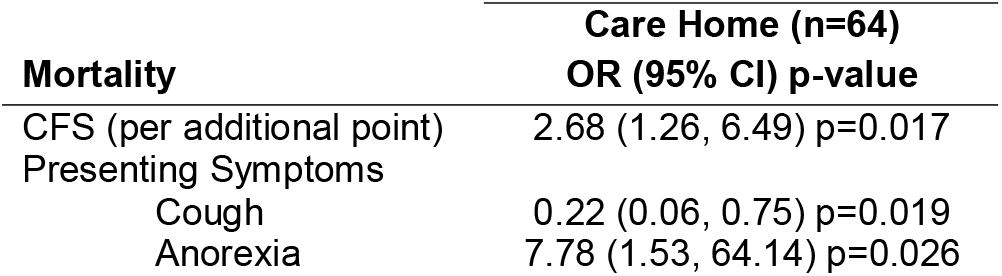
Multivariable Associations with Mortality in Care Home Residents. Odds ratios rounded to 2 decimal places. p values indicate significance of association between outcome and (level of) covariate. CFS = Clinical Frailty Scale, OR = odds ratio, 95% CI = 95% confidence interval, n = number of participants.

## 4. Discussion

This single-centre study found that inpatients over the age of 80 with PCR positive COVID-19 presented with not only cough, fever and dyspnoea, but also with a high proportion of hypoactive delirium, falls and anorexia. These trends were more pronounced in individuals presenting from residential or nursing care, who were less likely to present with cough, and more likely to present with delirium. Atypical presentations that featured none of the cardinal features of cough, dyspnoea and fever were common in patients presenting from institutional care. Individuals presenting from care homes were no more likely to die overall than those presenting from the community, though presentation with higher levels of frailty or anorexia was associated with increased mortality for both groups. Taken together our findings suggest a high prevalence of atypical symptoms in older adults, particularly institutionalised populations, and a risk of increased mortality for those with less classic presentations.

Our study has several strengths. Firstly, the data were collected contemporaneously as part of routine clinical work, allowing for easy characterisation of presenting complaints and patient histories. To accurately categorise delirium, delirium subtype, CFS and presentation with falls or anorexia, geriatric syndromes were independently corroborated by a full review of medical records by two consultant geriatricians. Our data was collected from an area of the UK affected by high COVID-19 caseload early in the course of the UK pandemic. These data were also not without limitations. The comparisons drawn between institutional care and community care in our data do not account for differences between nursing home and residential home care, which cannot always be determined from address and admission history. Key differences may still exist between these subgroups. Likewise, community dwelling older adults have differing care needs, and homogenising this group risks ignoring those with high levels of social care including that provided by the private sector. However the higher trend in CFS displayed in those in institutional care suggests this was not a major issue within our sample. Using an endpoint of discharge versus death limits mortality analysis, as some of those discharged may subsequently die in the community; however this number is likely lower than would be expected in an older population, as few palliative discharges were made due to limitations on carer visits and return to residential care imposed by the infective nature of COVID-19. Likewise collected biomarker data from patients was sporadic in this population, limiting its use in models of mortality without extensive imputation.

Although the spectrum of symptoms seen here with COVID-19 presentation is in keeping with that seen in the general UK population [4], the high proportion of falls, delirium and anorexia suggests prior case reports [6-10] are indicative of a more widespread pattern. The suggestion that COVID-19 presents atypically more frequently in older adults, and particularly in care home residents, supports a prior study in a United States nursing home, where atypical symptoms alone were noted in just under 10% of PCR positive patients [12]. In particular the predominance of hypoactive delirium in this population is in keeping with response to critical illness in patients with pre-existing cognitive impairment [7].

Our findings are also consistent with previously reported associations between high mortality and older age [15, 18-20]. While we did not find associations between mortality and dyspnoea [21], cardiovascular disease [22] or male gender [23] reported elsewhere, our population represents severe disease admitted to secondary care with a high overall mortality of 65%. The high rates of cardiovascular disease and the slight weighting toward male gender in our cohort may be a marker of these trends. Associations seen here between frailty and increased mortality are similar to other studies exploring frailty in COVID-19 [24], though notably none of our cohort received mechanical ventilatory support. CFS score should not be the sole determinant of treatment suitability [25]. Of key interest are novel associations seen here between anorexia, cough and mortality, particularly in institutionalised populations.

Associations between increased mortality and anorexia might be explained by late presentation of those with COVID-19 to secondary medical services. Anorexia is a subtle sign, becoming apparent over a few days, and so might not be detected until further into the disease course. Likewise, those with cough might have been referred earlier given the prominence of cough in Public Health England (PHE) and WHO criteria for COVID-19 diagnosis [26], potentially explaining lower mortality rates in this group. Yet the contemporaneous lack of a reliable treatment for COVID-19, bar supportive measures, draws into question any mortality benefit of early versus late diagnosis. Acute kidney injury (AKI) has previously been found to be associated with increased mortality in COVID-19 [15, 18]. While this may be related to disease associated systemic inflammation or a pro-coagulable state, dehydration will no doubt exacerbate these issues. Anorexia may therefore exacerbate the risk or severity of AKI and its associated poor prognosis, and be amenable to supportive therapy. This area needs further research; it is important to highlight therefore that anorexia should not be considered as a reason for limiting secondary care access for care home residents.

Atypical presentations are a key feature of geriatric medicine, and their continued existence within the COVID-19 syndrome is in keeping with the complex nature of the specialty. Their association with institutional care and higher mortality is worth exploring further, and may not be wholly explained by the ramifications of delayed presentation. Given the lack of evidence for higher mortality in those admitted from care homes, vigilance in diagnosis is recommended within these settings; particularly where hypoactive delirium and anorexia are concerned. Even without established pharmacological therapy, early symptom recognition combined with isolation and supportive care may provide marked benefit for these vulnerable populations.

## Data Availability

Data is available on reasonable request.

## 5. Declarations

### 5.3 Availability of data and material

Data is available on reasonable request.

### 5.4 Code availability

All code was compiled using R Statistics version 3.6.3, and is available on reasonable request.

### 5.5 Authors’ contributions

MJR was responsible for data analysis with additional input from SEB. MJR & DLB devised the research question and wrote the first draft of the manuscript. SEB conceived and initiated the data collection project. All authors contributed to interpreting the data and writing the final paper.

## 5.6 Acknowledgements

The authors would like to thank Ezgi Ozcan, Tom Burns, Lisa Amani and Rabia Warraich for their assistance in collecting the original data, and Dr Dean Creer, Dr Hannah Jarvis and Dr Amina Jaffer for assistance with planning the database.

